# Association of University Reopening Policies with New Confirmed COVID-19 Cases in the United States

**DOI:** 10.1101/2020.12.11.20247353

**Authors:** Yang Li, Cheng Ma, Weijing Tang, Xuefei Zhang, Ji Zhu, Brahmajee K. Nallamothu

## Abstract

**Importance:** Reopening of universities in the U.S. has been controversial in the setting of the coronavirus disease 2019 (COVID-19) pandemic.

**Objective:** To investigate (1) the association between new COVID-19 cases since September 1^st^ with the number of students returning to campus in each county across the U.S. and (2) how different reopening policies at universities correlated with new COVID-19 cases.

**Design:** Observational cohort study using publicly available data sources. Multivariable regression models estimated both effects of university reopening and different reopening policies.

**Settings and Participants:** Populations in U.S. counties reporting new confirmed COVID-19 cases from August 1^st^ to October 22^nd^.

**Exposures:** (1) total enrollment of students under the in-person or hybrid policies per county population and (2) proportion of online and hybrid enrollment within each county.

**Main Outcomes and Measures:** Mean number of daily new confirmed COVID-19 cases per 10,000 county population from September 1^st^ to October 22^nd^.

**Results:** For 2,893 counties included in the study, mean number of daily confirmed cases per 10,000 county population rose from 1.51 from August 1^st^ to August 31^st^ to 1.98 from September 1^st^ to October 22^nd^. Mean number of students returning to universities was 2.1% (95% CI, 1.8% to 2.3%) of the county population. The number of students returning to campus had an increased association (β = 2.006, P < 0.001) with new confirmed COVID-19 cases within the local county region where the institution resided. For 1,069 U.S. counties with universities, the mean proportion of online enrollment within each county was 40.1% (95% CI, 37.4% to 42.8%), with most students enrolling in-person or hybrid mode. In comparison to holding classes in-person, reopening universities online (β = -0.329, P < 0.001) or in a hybrid mode (β = -0.272, P = 0.012) had a decreased association with new confirmed COVID-19 cases.

**Conclusions and Relevance:** A higher number of students returning to campus in U.S. counties was associated with an increase in new confirmed COVID-19 cases; reopening online or partially online was associated with slower spread of the virus, in comparison to in-person reopening.

**Key Points:** *Question:* Are students returning to universities and specific reopening policies associated with new confirmed coronavirus cases in United States?

*Findings:* In this cohort study of 2,893 U.S. counties, the number of students returning to campus was significantly associated with a higher number of new confirmed COVID-19 cases. In 1,069 U.S. counties with universities, online or hybrid reopening was significantly associated with a lower risk of new cases compared with in-person reopening.

*Meaning:* An increased risk of coronavirus infection was seen in surrounding regions after universities reopened last fall, and this effect was largest in those holding in-person classes.

## INTRODUCTION

The new 2020 academic year for universities and colleges in the U.S. began in the midst of the COVID-19 pandemic in late August and early September. During March and April, most schools moved their classes fully online and locked down campuses. For the 2020 fall semester, however, many schools opted to reopen fully or partially in-person, given a desire to optimize learning environments and the challenges of virtual classrooms. Other schools, however, chose to institute an online or hybrid mode of learning given concerns for student safety. Though all schools have tried to take many public health actions to ensure reopening occurs safely, prominent reports in the lay media suggest universities and colleges experienced outbreaks of COVID-19 cases after students returned that have extended to surrounding communities.^1,2^

Despite such anecdotal observations, the precise effects of students returning to campus on their local regions remain uncertain. Furthermore, how specific university and college reopening policies may be linked to the spread of coronavirus is still unclear. In this study, we explore these concerns more specifically. We focus both on the association between the spread of coronavirus since September 1^st^ with the number of students returning to campus in each county across the U.S. as well as how different reopening policies at universities are correlated with new confirmed COVID-19 cases.

## MATERIALS AND METHODS

### Study Design

Our study had 2 key aims. First, we looked at all U.S. counties to study the association between the spread of coronavirus from August 1^st^ to August 31^st^ with the number of students returning to campus. In this aim of the study, we did not distinguish between different reopening policies of universities and colleges (except for excluding those with an online mode of reopening only) and primarily focus on the proportion of population enrollment in universities and colleges in the local county region to understand how the number of students returning to campus may be related to new confirmed COVID-19 cases more generally in their community. Second, we narrowed our focus to only those U.S. counties with universities and colleges to specifically study the association between different reopening policies and new confirmed COVID-19 cases. Using publicly available information,^3,4^ we divided different policies into three categories: in-person, hybrid, and online. We then examined the extent to which virtual classes through hybrid or online reopening – rather than in-person classes – was associated with new confirmed COVID-19 cases.

### Data Source

We aggregated multiple publicly available datasets for these analyses. First, we used county-level daily new confirmed cases during the period of August 1^st^ to October 22^nd^, 2020 (the specific dates of our study time period) from the COVID-19 tracking project^5^ by 1Point3Acres^6^ which was last updated on October 23^rd^, 2020. Second, we obtained state-level cumulative testing rates from the CDC COVID Data Tracker.^7^ Third, we collected various university and college reopening policies and data on their enrollment from 2,958 institutions across 1,253 counties utilizing resources from the Chronicle of Higher Education^3^, which was last updated on October 1^st^, 2020. (Note: when we use the term university in isolation below we refer to both universities and colleges of higher education with learners that enroll after secondary school.) These data were originally provided by the College Crisis Initiative at Davidson College.^4^ Fourth, we obtained county-level demographics data and land area from the United States Census Bureau^8^ and socioeconomic data from the Economic Research Service in United States Department of Agriculture.^9^ Socioeconomic data included the county-level resident population estimates by age and race in 2019, the county-level median household income in 2018, and the county land area in square miles in 2010.

Our study design focused on analyses done at the county level. The COVID-19 tracking project dataset initially provided county-level daily confirmed cases for 3,086 counties (out of 3,141 counties in the U.S.). To focus on in-person, online, and hybrid policies of interest, we excluded 163 counties that contained any university with “other” or “undetermined” policies given uncertainty as to whether students were returning to campus. We also observed a handful of “negative” daily confirmed cases (i.e., the number of COVID-19 cases was less than zero). On investigation, we found these negative cases were used to correct historical mistakes in the county-level data (i.e., a case was re-categorized the following day from being positive to negative). Given concerns that such data from counties with large numbers of corrections are likely to be unstable and may indicate limited surveillance systems, we excluded 30 (∼1%) counties that contained at least one day with more than 5 corrected cases during our study period. This resulted in 2,893 counties in our final analysis.

### Outcome Variables

For both aims, our main outcome variable was the mean number of daily new confirmed COVID-19 cases per 10,000 county population from September 1^st^ to October 22^nd^ at the county level. We used the starting date of September 1^st^ to capture the trend of confirmed cases after reopening. Most universities reopened near late August or early September, and we felt this would capture an adequate period of time of exposure after students returned back to campus. Moreover, we considered the mean number of daily new confirmed cases over a relatively long period (until the end date of analysis on October 22^nd^, 2020), rather than shorter intervals, to smooth out the weekly periodic pattern and a potential lag in impact of various reopening policies. Finally, we standardized the county-level average number of daily new confirmed cases by the county population (per 10,000) as the total population varies significantly over different counties.

### Exposure Variables

We used different exposure variables for the two aims. To study the impact of universities’ reopening, we focused on the number of students enrolled through in-person or hybrid classes trying to examine the effect of reopening between counties with and without universities. To further compare different reopening policies, we focused on specific reopening policies in only counties with universities. The original dataset for reopening policies mainly categorized five policies including “fully in person”, “primarily in person”, “hybrid”, “primarily online”, and “fully online”. For ease of interpretation, we further merge “fully in person” and “primarily in person” as in-person policy, and “fully online” and “primarily online” as online policy, which resulted in three policy categories: in-person, online, and hybrid in our analysis. Below we describe the exposure variables used for the two aims in detail.

To study the impact of universities’ reopening, we used an exposure variable that captured the number of university students returning to their campus within each county. Given that the enrollment and policies of universities in each county may differ when more than one university is present in a county, we defined the exposure variable for this county as follows. We first aggregated all universities in a county and summed up the enrollment of students under the in-person or hybrid policies (where students would likely return to campus). Then the exposure variable was obtained as the ratio of this sum and the total county population (*ENROLLP*). Note that we used the total enrollment of universities under the above two policies as a reasonable proxy of the number of university students returning to the campus. And, as there is a large variance among universities’ enrollment, the proposed exposure variable better characterizes the population flow, in comparison to alternatives such as the number of universities with in-person or hybrid policies. A higher value of the exposure variable means that more students stayed in physical buildings and were involved in onsite campus activities per county population. The exposure variable equals zero if there was no university in a region (or all universities in a region took online reopening policy).

To compare different reopening policies across universities, we focused on a subset of 1,069 counties with universities. The main exposure variables were the proportion of enrollment for three policies within each county (*ONLINE%, HYBRID%, INPERSON%*). For example, for each county, the “in-person” variable was the ratio of the total enrollment of all universities taking in-person policy in this county to the total county enrollment. The sum of three policy exposure variables equals to one. We were interested in whether the proportion of hybrid and online enrollment would be negatively associated with the COVID-19 confirmed cases when taking in-person enrollment as the baseline. We showed an example of this calculation in Figure 4 for Washtenaw County where the University of Michigan is one of several schools in the region.

### Other Control Variables

We considered the following control variables in models for both aims. We first included the mean number of daily new confirmed COVID-19 cases per 10,000 county population from August 1^st^ to August 31^st^ (*AUGP*). Examining new confirmed COVID-19 cases in August before reopening occurred would allow us to account for baseline differences in the severity of coronavirus infections across counties. We also controlled for five county-level demographic and economic characteristics that could potentially be associated with new confirmed COVID-19 cases including: 1) the proportion of population aged over 65 years (*AGE65*), 2) the proportion of population aged below 20 years (*AGE20*), 3) the proportion of population of racial minorities (non-White) (*RACEMINO*), 4) the median household income as of 2018 (*MEDHHINC*), and 5) COVID-19 testing rates at the state level (*TR*). Note that for the variable testing rates, we only had data available at the state level and used this as a proxy for county-level resources for testing.

### Statistical Analysis

We used multivariable linear regression models to estimate the effects of universities’ reopening and different reopening policies. Note that the enrollment proportion of three policies sum up to 1, so we dropped the covariate in-person enrollment proportion to avoid the issue of collinearity among covariates. We used a p-value of less than 0.05 to indicate statistically significant evidence of an association between the covariate and the outcome variable.

### Lastly, we conducted three sensitivity analyses

First, we conducted another analysis to compare the early-stage (within a month) and late-stage (after a month) impact of universities’ reopening. Specifically, for each county, we considered two periods of time, one in September and the other in October. Thus, each county has two outcome measurements, corresponding to the mean number of daily new confirmed cases per 10,000 county population in September and in October respectively. We also constructed an indicator variable indicating whether the period under consideration is October (*I(Oct)*), and added itself and its interactions with the mean daily new confirmed proportion in August and the enrollment proportion into the linear regression model to account for the variation over time.

Second, we changed the exposure variable - the in-person and hybrid enrollment proportion - into a binary variable. This variable is 1 if the in-person and hybrid enrollment proportion is greater than zero, and is 0 otherwise. The goal of this sensitivity analysis was to determine if a “dose-response” effect was noted based on the proportion of in-person and hybrid enrollment (rather than its mere presence in a county).

Third, because the impact of universities’ reopening to counties with large populations can be relatively minor, we further removed 87 counties with the top 5% total county population and re-estimated the model based on this cohort.

## RESULTS

### Cohort Characteristics

Our final study cohort included 2,893 U.S. counties with a subset of 1,069 counties where at least one university or college was present. Characteristics of these U.S. counties are provided in Table 1. The mean of daily confirmed cases per 10,000 county population was 1.51 from August 1^st^ to August 31^st^, and 1.98 from September 1^st^ to October 22^nd^. The overall total number of daily new confirmed cases across the U.S. shows an increasing trend starting from early September (Figure 1). Mean number of students returning to schools was 2.1% (95% CI, 1.8% to 2.3%) of the county population. For those counties with universities, the proportion of online enrollment within each county is on average 40.1% (95% CI, 37.4% to 42.8%), with the rest of the students enrolling in-person or hybrid mode.

**Table 1.** Cohort Characteristics. The outcome is SEPTP (the average number of daily new confirmed COVID-19 cases per 10,000 county population after September 1^st^). The main exposure variable is ENROLLP (in-person + hybrid enrollment proportion). The control variables are: AUGP (the average number of daily new confirmed COVID-19 cases per 10,000 county population in August), AGE20 (the proportion of young people under the age of 20), AGE65 (proportion of older adults 65 years or older), RACEMINO (the proportion of population of racial minorities, i.e., non-White), MEDHHINC (the median household income as of 2018), TR (the COVID-19 testing rates at the state level).

|  | **All 2,893 counties**<br>**under study** | **1,069 counties with**<br>**colleges** |
| --- | --- | --- |
| **Variables** | **Mean(sd)** | **Mean(sd)** |
| SEPTP | 1.98(1.66) | 1.87(1.45) |
| AUGP | 1.51(1.78) | 1.42(1.17) |
| ENROLLP | 2.05(5.79) | N/A |
| INPERSON% | N/A | 36.1(45.1) |
| HYBRID% | N/A | 23.8(38.8) |
| ONLINE% | N/A | 40.1(45.1) |
| AGE20 | 24.3(3.52) | 24.8(3.00) |
| AGE65 | 19.9(4.70) | 18.2(3.82) |
| RACEMINO | 15.1(15.9) | 17.6(1.59) |
| MEDHHINC<br>(thousand USD) | 52.2(13.5) | 55.0(14.4) |
| TR | 0.413(0.146) | 0.422(0.144) |

**Figure 1.**
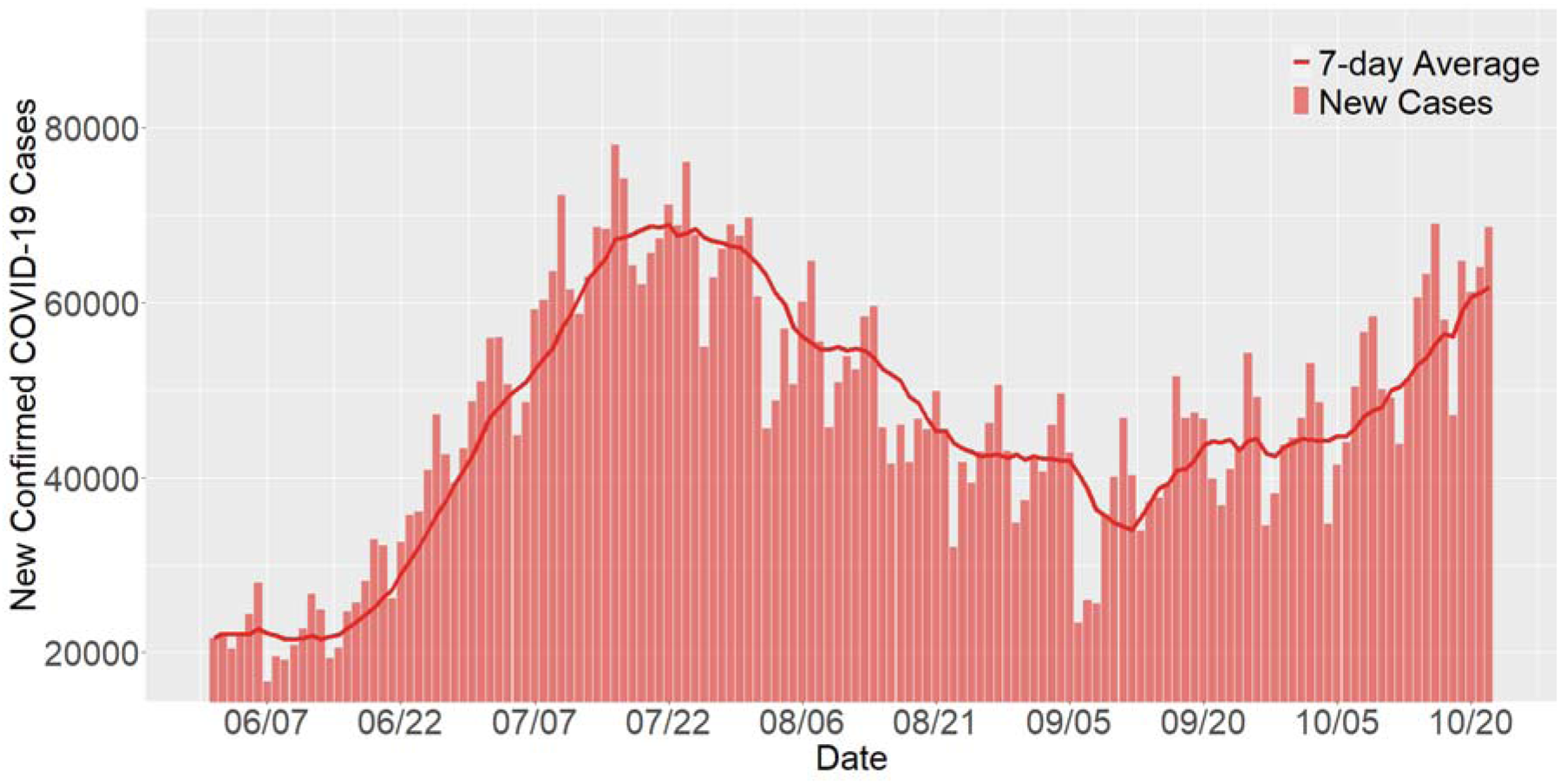
Nationwide total number of daily new confirmed COVID-19 cases since June 1^st^ and their 7-day average. The number of daily new confirmed COVID-19 cases shows an increasing trend starting from early September that coincides with the time of school reopening.

### Students Returning to Campus and New Confirmed COVID-19 Cases

Mean number of daily new confirmed COVID-19 cases per 10,000 county population from June 1^st^ to October 22^nd^ over U.S. counties stratified by the presence of returning students is plotted in Figure 2. Mean number of daily new confirmed COVID-19 cases in U.S. counties with the top 50% in-person or hybrid university enrollment rates increased at a higher rate compared with U.S. counties without in-person or hybrid universities. This separation appeared largest during an early phase in September with infections rising in U.S. counties without in-person or hybrid colleges in October.

**Figure 2.**
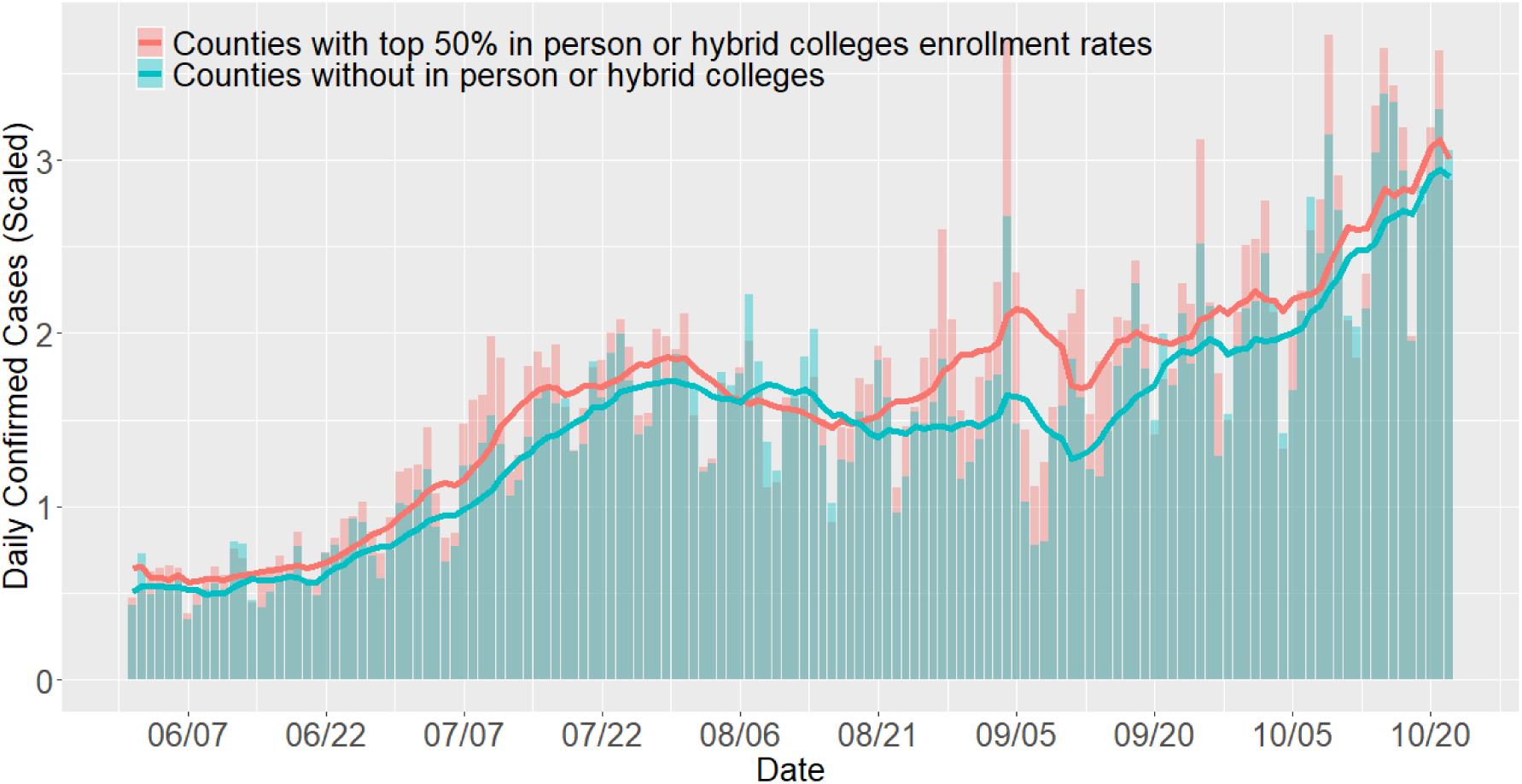
The mean number of daily new confirmed COVID-19 cases per 10,000 county population over U.S. counties without universities with in-person or hybrid reopening policies, and U.S. counties with the top 50% in-person or hybrid university enrollments per 10,000 county population from June 1^st^ to October 22^nd^. The solid curves represent their 7-day average. The mean number of daily new confirmed COVID-19 cases in U.S. counties with the top 50% in-person or hybrid college enrollment rates appears to increase as compared with U.S. counties without in-person or hybrid colleges. Interestingly, this separation appears to be greatest during an early phase in September with infections rising in U.S. counties without in-person or hybrid colleges in October.

The result of our fitted model after multivariable adjustment is shown in Table 2. Counties with higher proportions of in-person or hybrid university enrollment were associated with higher confirmed case rates since September (β = 2.006, p-value < 0.001). That means, for every 10% rise in the ratio of in-person or hybrid university enrollment to the county population, the mean daily new confirmed cases would increase by about 0.2 persons per 10,000 county population (e.g., 41.8 cases per 30 days in a median-size U.S. county where at least one university was present).

**Table 2.** The estimated coefficients on 2,893 U.S. counties with or without universities and colleges. The main exposure variable is the sum of in-person and hybrid enrollment proportion (ENROLLP). The control variables are the same as Table 1

|  | **Estimate** | **Std Error** | **P-value** |
| --- | --- | --- | --- |
| Intercept | -2.507 | 0.495 | <0.001 |
| AUGP | 0.204 | 0.017 | <0.001 |
| AGE20 | 14.148 | 1.134 | <0.001 |
| AGE65 | 4.373 | 0.954 | <0.001 |
| ENROLLP | 2.006 | 0.508 | <0.001 |
| RACEMINO | -0.955 | 0.199 | <0.001 |
| MEDHHINC<br>(thousand USD) | -0.015 | 0.002 | <0.001 |
| TR | 1.810 | 0.201 | <0.001 |

Additional variables beyond students returning to campus were associated with new confirmed COVID-19 cases as well. For example, the average confirmed case rate in August was positively correlated with the confirmed case rate after September (β = 0.204, p-value < 0.001). The proportion of young people under the age of 20 and the proportion of older adults 65 years or older also were both positively associated with the outcome (p-values < 0.001). This could be due to the fact that older adults are at higher risk of getting COVID-19, and young people would likely be going back to school after September, bringing higher chances of COVID-19 spread. Median household income, which can in general represent the local economic status, was negatively correlated with the outcome (β = -0.015, p-value < 0.001) suggesting lower numbers of new confirmed COVID-19 cases in wealthier regions. As expected, the regional testing rate was positively associated with the outcome (β = 1.810, p-value < 0.001). Finally, we have found that the proportion of population of racial minorities in a county was negatively correlated with the outcome (β = -0.955, p-value <0.001). This result was robust using other control variables that represent racial minorities (e.g., African American proportion in a population). This may be due to our evaluation of new confirmed cases late in the pandemic (i.e., time after September), instead of the whole pandemic period where rates of infection have been higher in U.S. counties with higher proportions of minority populations.

### Reopening Policies and New Confirmed COVID-19 Cases

Figure 3 shows the mean number of daily confirmed COVID-19 cases per 10,000 population over 318 U.S. counties with all universities taking an in-person policy, 173 U.S. counties with all universities taking a hybrid policy, and 303 U.S. counties with all universities taking an online policy. Daily confirmed cases per 10,000 population of three groups were similar until the end of August, but after that, there were gaps between 3 groups (as reopening of schools occurred). Among 1,069 counties with universities, we estimated models after multivariable adjustment and displayed these results in Table 3. Online enrollment proportion was negatively associated with the outcome (β = -0.329, p-value < 0.001). This result could be interpreted in this way: fixing the hybrid enrollment proportion, if we move 10% of the enrolled university students in a county from in-person to online mode, the mean number of daily new confirmed cases per 10,000 county population would have decreased by approximately 6.85 cases per 30 days in a median-size U.S. county where at least one university was present. We also found the hybrid policy enrollment proportion was negatively correlated with the outcome (β = -0.272, p-value = 0.012). This suggests that moving 10% of the enrollment students from in-person mode to hybrid mode would reduce the mean number of daily new confirmed cases per 10,000 county population by approximately 5.67 cases per 30 days in a median-size U.S. county where at least one university was present. The interpretation of other control variables is similar to our earlier results.

**Table 3:** The estimated coefficients on 1,069 U.S. counties with universities and colleges. The main exposure variables are policies as enrollment percentages (ONLINE% and HYBRID%). The control variables are the same as Table 1.

|  | **Estimate** | **Std Error** | **P-value** |
| --- | --- | --- | --- |
| Intercept | -0.729 | 0.743 | 0.327 |
| AUGP | 0.425 | 0.036 | <0.001 |
| AGE20 | 12.109 | 1.824 | <0.001 |
| AGE65 | -1.511 | 1.499 | 0.314 |
| ONLINE% | -0.329 | 0.095 | <0.001 |
| HYBRID% | -0.272 | 0.108 | 0.012 |
| RACEMINO | -1.454 | 0.264 | <0.001 |
| MEDHHINC<br>(thousand USD) | -0.017 | 0.003 | <0.001 |
| TR | 1.581 | 0.267 | <0.001 |

**Figure 3.**
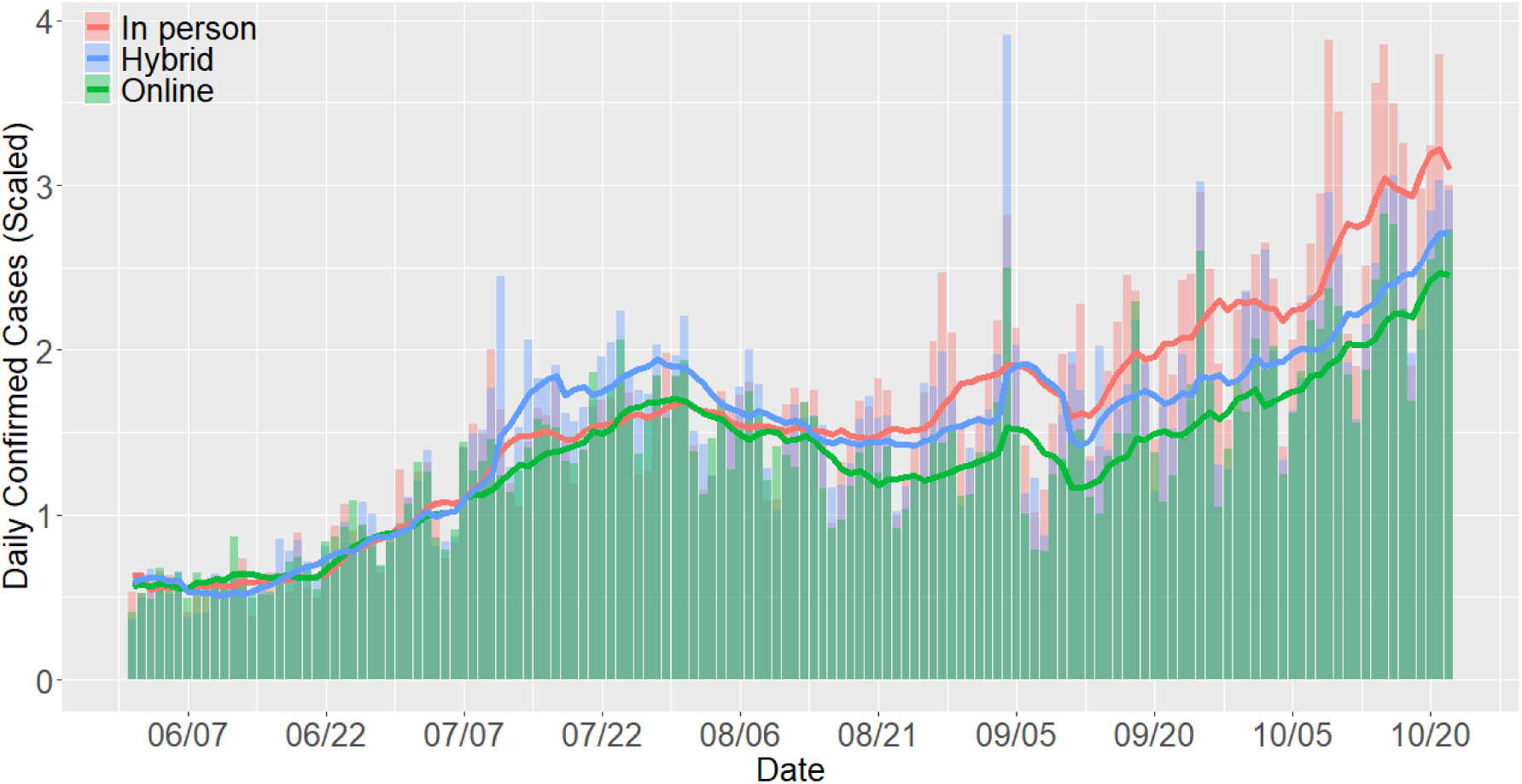
The mean number of daily confirmed COVID-19 cases per 10,000 population over 318 U.S. counties with universities taking an in-person policy, 173 U.S. counties with universities taking a hybrid policy, and 303 U.S. counties with universities taking an online policy. The solid curves represent their 7-day average.

**Figure 4.**
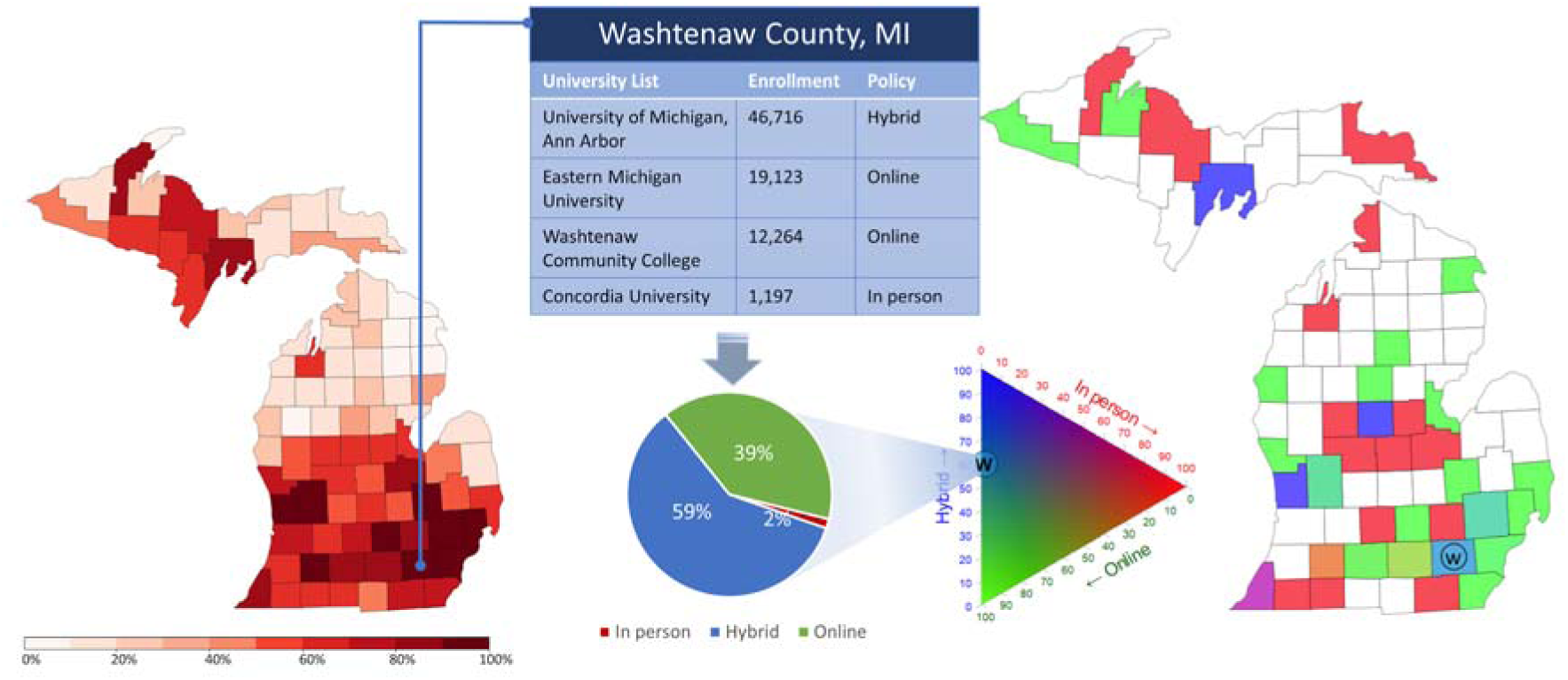
We illustrate how to calculate our exposure variables using our state of Michigan. Left: The percentile rank of the mean daily confirmed cases from September 1^st^ to October 22^nd^ relative to all counties in the U.S. Middle: Schematic illustration of how to construct the main exposure variables in our analysis of U.S. counties with universities and colleges: the proportion of enrollment for three policies within each county. Right: The geographic distribution of universities’ reopening policies over counties in Michigan.

### Sensitivity Analysis

We performed several sensitivity analyses. We found evidence of an early-period effect with enrollment proportion noted to be a significant variable in affecting the number of new confirmed COVID-19 cases in September, but with its association becoming insignificant in October (p-value = 0.054). This suggests that the effect of school reopening may depend on the time, and the effect is more important in the first few weeks after reopening with a “catch-up” phase in U.S. counties without universities. This time-varying effect is observed in Figure 2, where there was an initial case surge in September with a later rise in cases in October. Secondly, converting the continuous value of enrollment proportion into a binary one leads to weaker association between new confirmed COVID-19 cases and university enrollment. This implies the value of measuring the specific number of in-person and hybrid enrollment as a continuous variable, rather than just a simple indicator, to reflect the extent of university students returning to the campus. Finally, we performed a sensitivity analysis that excluded U.S. counties with large populations to ensure our findings were not susceptible to outliers; these results were consistent with our earlier findings. The details of these model fitting results are summarized in Supplementary Information.

## DISCUSSION

An outbreak of COVID-19 cases on college campuses has been reported since students returned to campus in the fall 2020 semester. A recent analysis performed by the *NY Times* suggested that a spike in cases occurred in U.S. counties with universities starting in September 2020, and that many universities have taken aggressive actions to prevent broader outbreaks as a result^10.^ In this study, we examined the effect of reopening in universities and colleges on the spread of COVID-19 in their surrounding communities during the fall after accounting for several additional features that may contribute to infections. Our findings could be summarized in two main points. First, we found that returning to campus was associated with an increase in new confirmed COVID-19 cases in the counties in which the universities and colleges reside. We estimated an additional 41.8 cases per 30 days in a median-size U.S. county where at least one university was present. Second, we assessed the effects of three different reopening policies and found that reopening online or partially online was associated with slower spread of the virus, in comparison to in-person reopening.

Our findings have important implications for understanding how the reopening of universities and colleges in the US may have affected infection rates of COVID-19 in their surrounding communities. They suggest that school reopening, especially in-person reopening, brought higher risks to their neighboring areas. Follow-up data on the winter/spring 2021 semester may help to elucidate these findings further, although regions with universities may already be paying closer attention to possible resurgent outbreaks. This includes effective actions such as university-wide screening, contact tracing, enforcing wearing masks indoors and strict social distancing, flexible making-up classes policies, etc.^11^ Taiwan reported low rates of infection in their communities using such policies, however, this was in the context of societal-wide adoption of strict infection control practices.^12^

These findings should be interpreted in the context of the following limitations. First, we studied the effects of university reopening policies using observational data with multivariable adjustment. The choice of control variables may not be a comprehensive list, and residual confounding from other sources of variability in county-level rates of COVID-19 cases is possible. However, we did adjust for several key factors, and our results were stable over sensitivity analyses. Second, the actual reopening dates may vary among universities, ranging from late August to mid-September. In our analysis, the calculation of outcome variable, which is based on the cutoff date September 1^st^, could be viewed as approximation of confirmed rates after university reopening. Not only does the start vary across different campuses, but even when students arrive back on campus it can be difficult to ascertain directly their time of contact with others. Finally, testing capacity and the accuracy of reported numbers of tests and cases might have varied across counties or over time as the COVID-19 pandemic developed. This issue could have also introduced some noise to the measurement we used of new confirmed COVID-19 cases.

In conclusion, we found a higher number of new confirmed COVID-19 cases in U.S. counties after students returned to universities and colleges during the fall of 2020. Among U.S. counties with universities and colleges, reopening with in-person classes was associated with a higher number of new cases compared with holding online classes or a hybrid mode of learning. These findings suggest that students returning to campus increased the risk of coronavirus infection while providing evidence supporting potential mitigation strategies such as online learning.

## Data Availability

All datasets used are publicly available. Our analysis was conducted using R statistical software version 3.6.0 and 3.6.1. All statistical code is available for replication at https://github.com/yli15/COVID-19-research.

https://github.com/yli15/COVID-19-research.git.

## ACKNOWLEDGEMENTS

This work was done through support and access to the American Heart Association COVID-19 Data Challenge Award and its Precision Medicine Platform. Our team was awarded the final prize as part of this competition. We are grateful to Ms. Jessica Baker with her assistance in preparation of this manuscript.

