## Supplemental Information for "Association of University Reopening Policies with New Confirmed COVID-19 Cases in the United States"

*Brahmajee K. Nallamothu, 132-W, B16, 2800 Plymouth Road, Ann Arbor, Michigan 48109.

**Phone Number:** (734)647-1624

**This PDF file includes:**

Supplementary text

Tables S1 to S4

Supplementary Information Text

**Sensitivity Analysis.**

Comparison of the early-stage and late-stage impact of universities’ reopening (for Aim 1 only)

We studied the effect of enrollment proportion on the outcome in September and October separately. We found that the effect of university enrollment proportion has a more significant impact on increasing the risk of COVID-19 spread in September, which is right after reopening. Such effects become insignificant in October (p-value: 0.054). Our main study covers both September and October and captures the significant effect of such variable. The result of the fitted model is shown in Table S1.

Binarizing the covariate in-person and hybrid enrollment proportion (for Aim 1 only)

The covariate binarized enrollment indicator (i.e., whether a county has any student enrolled in in-person or hybrid mode) was not significant (p-value: 0.93). This demonstrates the necessity of measuring the exact number of in-person and hybrid enrollment as a continuous variable, rather than just a simple indicator, to reflect the extent of university students returning to the campus within each county. The result of the fitted model is shown in Table S2.

Excluding large population counties

We performed our analysis for Aim1 and Aim 2 after filtering the top 5% largest counties based on county population. The re-estimated coefficients are very similar to the original model, which suggests that our analysis is not sensitive to including counties with large populations. The result of the fitted model is shown in Table S3 and Table S4.

Table S1. Sensitivity analysis: comparison of the early-stage and late-stage impact of universities’ reopening for Aim 1. I(Sep) is the September indicator, and I(Oct) is the October indicator, and other variables are the same as Table 1.

|  | **Estimate** | **Std Error** | **P-value** |
| --- | --- | --- | --- |
| Intercept | -3.493 | 0.445 | <0.001 |
| I(Oct) | 1.389 | 0.069 | <0.001 |
| AUGP | 0.404 | 0.021 | <0.001 |
| AGE20 | 15.263 | 1.016 | <0.001 |
| AGE65 | 4.854 | 0.855 | <0.001 |
| RACEMINO | -0.989 | 0.179 | <0.001 |
| MEDHHINC | 0.016 | 0.002 | <0.001 |
| TR | 1.969 | 0.180 | <0.001 |
| AUGP * I(Oct) | -0.460 | 0.029 | <0.001 |
| ENROLLP * I(Sep) | 2.637 | 0.635 | <0.001 |
| ENROLLP * I(Oct) | 1.224 | 0.635 | 0.054 |

**Table S2**. Sensitivity analysis: binarizing enrollment proportion (in-person + hybrid) for Aim 1. ENROLL_BINARY means enrollment indicator, and other variables are the same as Table 1.

|  | **Estimate** | **Std Error** | **P-value** |
| --- | --- | --- | --- |
| Intercept | -2.146 | 0.495 | <0.001 |
| AUGP | 0.202 | 0.017 | <0.001 |
| AGE20 | 13.793 | 1.139 | <0.001 |
| AGE65 | 3.553 | 0.957 | <0.001 |
| ENROLL_BINARY | -0.006 | 0.067 | 0.93 |
| RACEMINO | -0.993 | 0.2 | <0.001 |
| MEDHHINC | -0.016 | 0.002 | <0.001 |
| TR | 1.811 | 0.202 | <0.001 |

**Table S3.** Sensitivity analysis: excluding large population counties for Aim 1. All variables are the same as Table 2.

|  | **Estimate** | **Std Error** | **P-value** |
| --- | --- | --- | --- |
| Intercept | -2.542 | 0.505 | <0.001 |
| AUGP | 0.203 | 0.017 | <0.001 |
| AGE20 | 14.259 | 1.156 | <0.001 |
| AGE65 | 4.271 | 0.974 | <0.001 |
| ENROLLP | 1.971 | 0.515 | <0.001 |
| RACEMINO | -0.926 | 0.206 | <0.001 |
| MEDHHINC | -0.015 | 0.003 | <0.001 |
| TR | 1.852 | 0.206 | <0.001 |

Table S4. Sensitivity analysis: excluding large population counties for Aim 2. All variables are the same as Table 2.

|  | **Estimate** | **Std Error** | **P-value** |
| --- | --- | --- | --- |
| Intercept | -0.944 | 0.783 | 0.228 |
| AUGP | 0.429 | 0.038 | <0.001 |
| AGE20 | 13.176 | 1.929 | <0.001 |
| AGE65 | -1.787 | 1.585 | 0.26 |
| ONLINE% | -0.285 | 0.099 | 0.004 |
| HYBRID% | -0.253 | 0.112 | 0.024 |
| RACEMINO | -1.529 | 0.284 | <0.001 |
| MEDHHINC | -0.018 | 0.003 | <0.001 |
| TR | 1.658 | 0.284 | <0.001 |

1. †: These authors contributed equally to this work.
